# Single and 2-dose vaccinations with MVA-BN^®^ induce durable B cell memory responses in healthy volunteers that are comparable to older generation replicating smallpox vaccines

**DOI:** 10.1101/2022.09.07.22279689

**Authors:** Heiko Ilchmann, Nathaly Samy, Daniela Reichhardt, Darja Schmidt, Jacqueline D Powell, Thomas PH Meyer, Günter Silbernagl, Rick Nichols, Heinz Weidenthaler, Laurence De Moerlooze, Liddy Chen, Paul Chaplin

## Abstract

While the MVA-BN vaccine has been proven protective against smallpox and monkeypox, the long-term immunological persistence or booster effect has not been described. In this set of clinical studies, participants who had never been immunized against smallpox were randomized to receive, 4 weeks apart: 2 placebo vaccinations (PBO group, N =181); 1 MVA-BN vaccination followed by placebo(1×MVA group, N =181); or 2 MVA-BN vaccinations (2×MVA group, N = 183). In addition, participants with a history of smallpox vaccination received 1 MVA-BN booster (HSPX^+^ group, N = 200). The 1×MVA and 2×MVA groups responded with increases in neutralizing antibody (nAb) GMTs at Week 2 (5.1 and 4.8, respectively) that further increased at Week 4 (7.2 and 7.5). Two weeks after the second primary vaccination in the 2×MVA group (at Week 6), nAb GMT peaked (45.6) before stabilizing 2 weeks thereafter (at Week 8) (34.0). In the HSPX^+^ group, a rapid anamnestic response was observed with a peak nAb GMT at Week 2 (175.1) that was much larger than the peak responses in either of the primary vaccination (1× or 2×MVA) dose groups of smallpox vaccine-naïve subjects. Persistence of nAbs relative to baseline was observed at 6 months in all groups (highest in HSPX^+^), with a return to near baseline nAb levels 2 years later. Subsets of ∼75 participants each, who received primary vaccinations in the 1×MVA and 2×MVA groups, were administered an MVA-BN booster 2 years later. Both booster dose (BD) groups exhibited rapid anamnestic responses with nAb GMTs that peaked 2 weeks post-booster (80.7 and 125.3). These post-booster titers in the 1×MVA and 2×MVA groups were higher than those observed at any timepoint following primary vaccination, were comparable to HSPX^+^ subjects who had been administered a booster, and remained elevated at 6 months post-booster (25.6 and 49.3). The observed anamnestic responses, in the absence of sustained detectable nAbs, support the presence of durable immunological memory following MVA-BN immunization. No safety concerns were identified, and the most common adverse event following the 2-year MVA-BN booster was injection site erythema in 82.2% of participants.

**Clinical Trial Registry Numbers:** NCT00316524 and NCT00686582

**Highlights:**

- MVA-BN booster-induced anamnestic responses support durable immune memory
- One or two primary MVA-BN vaccinations induce similar durable B cell memory responses
- Anamnestic responses were observed in those immunized with MVA-BN 2 years earlier
- No safety concerns were revealed following a 2-year MVA-BN booster

## 1. Introduction

The discontinuation of smallpox vaccinations in the 1970s has left an increasing proportion of the world’s population susceptible to variola, the causative agent of smallpox, as well as to other orthopox viruses for which the vaccine provided cross-protection [1]. Thus, while the heightened risk of smallpox through occupational exposure [2, 3] or bioterrorism [4] remain concerning, the most prevalent human orthopox virus infection is currently monkeypox. In the 1970s and 1980s, cases of monkeypox were initially documented in west and central Africa, and infection was suspected to be primarily from animal to human with limited secondary spread between humans [5, 6]. Yet, over the past few decades, an increase in human monkeypox infection and transmission has been reported in Africa [7-9]. Beginning in 2018, isolated travelers from Nigeria experienced documented cases of monkeypox upon arriving in Israel, Singapore, and the UK, with secondary infection of some of these travelers’ close contacts serving as an early warning of the possible spread of monkeypox outside endemic regions [10, 11]. Since May 2022, a more geographically widespread monkeypox outbreak has been identified, primarily in men who identify themselves as gay, bisexual, or men having sex with men, leading to the declaration of a Public Health Emergency of International Concern by the World Health Organization [12]. These outbreaks highlight the absence of population protection against orthopox viruses, such as the monkeypox virus, and the urgency to better understand the priming capability of different schedules of protective vaccines.

Although world-wide protection against orthopox viruses was achieved with replicating vaccinia-based vaccines, these vaccines are associated with rare, but potentially serious, adverse reactions that include progressive vaccinia, eczema vaccinatum, generalized vaccinia, post-vaccinal encephalitis, and myopericarditis [13-16]. Risks of these adverse reactions are further increased in those who are immunocompromised or have exfoliative skin conditions [17-21]. These safety concerns have been addressed by the use of the highly attenuated modified vaccinia Ankara (MVA) virus to develop the non-replicating orthopox MVA-BN^®^ vaccine [22], which is substantially less reactogenic than traditional replicating smallpox vaccines [23, 24]. In the healthy adult population, MVA-BN is well tolerated [25-32] and induces immune responses comparable to traditional smallpox vaccines [23, 24, 33]. Similarly, no safety concerns with MVA have been identified in those living with human immunodeficiency virus [34-36], patients having received hematopoietic stem cell transplants [37], or people with exfoliative skin conditions [38, 39]. Based on these data, which are further supported by nonclinical studies demonstrating the protective efficacy of MVA-BN against monkeypox in nonhuman primates [40], MVA-BN has been approved for prevention of smallpox and monkeypox under the trade name JYNNEOS in the US [41], IMVANEX in Europe and UK [42], and IMVAMUNE in Canada [43].

The initial and follow-up clinical studies described herein were conducted to assess the safety, immunogenicity, and durability of MVA-BN in a mixed population, comprised of younger individuals who had never been immunized against smallpox and older individuals who were vaccinated against smallpox in the distant past using live-replicating vaccinia vaccines. The safety results of the initial study were previously reported [29] and are consistent with other studies demonstrating the favorable safety profile of MVA-BN, without an increased risk of myo-/pericarditis.

During the initial study, immune responses of those receiving either 1 or 2 primary vaccinations with MVA-BN were compared to anamnestic immune responses observed after an MVA-BN booster was given to participants immunized against smallpox in the distant past. Two years later, in a follow-up study, antibody persistence was investigated, and an MVA-BN booster was administered to a subset of participants who had received the 1- or 2-dose primary MVA-BN vaccinations in the initial study. Together these studies provide a comprehensive assessment of the duration and nature of immunological memory following different kinds of primary smallpox vaccinations that were either administered in the distant past with replicating vaccines or 2 years earlier with the nonreplicating MVA-BN vaccine. These findings are of particular relevance when defining optimal vaccination strategies following outbreaks in a supply-constrained environment and will also inform prophylaxis for those at increased risk of orthopox exposure, such as healthcare providers and military personnel.

## 2. Materials and Methods

### 2.1 Study design

Two clinical studies were conducted to assess safety, immunogenicity, and boostability of MVA-BN in healthy adult individuals with and without a history of smallpox vaccination. These studies were conducted at a single European site between 2006 and 2009.

The initial study (NCT00316524) was a phase 2, partially randomized, partially double blind, placebo-controlled, non-inferiority trial. All study participants without a history of smallpox vaccination (HSPX^−^) were randomized 1:1:1 to receive either 1 dose of MVA-BN followed by 1 dose of placebo 4 weeks later (1×MVA group), 2 doses of MVA-BN (2×MVA group) or 2 doses of placebo (PBO group). All participants with a history of smallpox vaccination (HSPX^+^) were assigned to receive a single booster dose of MVA-BN. The primary immunogenicity objective of this study was to compare the humoral immune responses of the HSPX^+^ and 2×MVA groups to determine if a single booster dose of MVA-BN in a previously vaccinated population could induce a response comparable to that induced by 2 MVA-BN primary vaccinations in a population who had never been vaccinated against smallpox. The primary safety objective of the study was to compare electrocardiogram (ECG) changes and cardiac symptoms across vaccination groups. Secondary objectives included comparisons of immune responses across all dose groups, an assessment of immune response kinetics following 1 or 2 doses of MVA-BN in HSPX^−^ participants, and a comparison of safety between HSPX^−^ participants in the 1×MVA and 2×MVA dose groups with those in the PBO group.

The follow-up study (NCT00686582) was a phase 2 open-label trial that rescreened participants from the 1×MVA, 2×MVA, and HSPX^+^ groups who participated in the initial study. Two years following their last dose in the initial study, immunogenicity assessments to assess antibody persistence were performed on participants from these 3 groups. An MVA-BN booster vaccination was also administered to a subset of those who had received either 1 or 2 primary doses of MVA-BN in the initial study, comprising the 1×MVA booster dose (BD) and 2×MVA BD groups. The primary objective was to evaluate the immune response elicited by the MVA-BN booster vaccination in the 1×MVA BD and 2×MVA BD groups. The secondary objectives were to evaluate long-term immunogenicity and safety.

### 2.2 Participants

All study-related procedures were in accordance with the provisions of the Declaration of Helsinki (2000/2008). All participants were informed verbally and in writing about the purpose, procedures, and potential benefits and risks and signed informed consent forms prior to participation in any study-related assessments.

For the initial study, men and nonpregnant women between 18 and 55 years of age were included if they had general lab values within normal limits at screening and agreed to use appropriate birth control measures. For the 1×MVA, 2×MVA, and PBO groups, participants were to have no known or suspected previous smallpox vaccination and no detectable vaccinia scar. For the HSPX^+^ group, participants were to have a known history of smallpox vaccination that was either documented or manifested as a typical vaccinia scar. The most recent previous smallpox vaccination must have been at least 5 years ago. Excluded from the study were participants with an uncontrolled serious infection; a history of or actively ongoing serious medical condition, including autoimmune disease or malignancy; impaired immunologic function, a history of organ transplantation, or chronic systemic use of immunosuppressant drugs anytime during the 6 months leading up to study involvement; a history of, or ongoing cardiovascular disease; an immediate family member with onset of ischemic heart disease before 50 years of age; or a history of intravenous drug abuse.

Consenting participants who had completed the initial study as part of the 1×MVA, 2×MVA, or HSPX^+^ groups were given the opportunity to have the persistence of their humoral response evaluated 2 years later, through an antibody assessment conducted at the beginning of the follow-up study. Participants originally randomized to the 1×MVA and 2×MVA groups in the initial study were eligible to receive a booster in the follow-up study if they had no major protocol deviations in the initial study, no general lab values outside normal limits, and no clinically significant ECG findings. Only the first 75 eligible participants in each of the 1×MVA and 2×MVA groups were planned to receive the booster vaccination.

### 2.3 Vaccine

MVA-BN is a highly attenuated, purified live vaccine [22], that together with the placebo was produced at IDT Biologika GmbH (Dessau-Rosslau, Germany) according to cGMP standards. The vaccine (initial study batch number: 170505; follow-up study batch number: 0040707) was provided in liquid frozen 0.5 mL aliquots and had a virus titer of ≥ 0.5 × 10^8^ TCID_50_ MVA. Each dose of placebo contained Tris Buffer, comprised of 1.21 g/L Tris, 8.526 g/L sodium chloride, 19.2 g/L dextran FP-40, 45.2 g/L sucrose, and 0.108 g/L L-glutamic acid mono-potassium salt at a pH of 7.5. Study vaccines and placebo (Tris Buffer) were shipped and stored at -20°C ± 5°C, avoiding direct light. All doses were administered subcutaneously with a 24- or 25-gauge needle in the upper arm according to standard clinical practice.

### 2.4 Immunogenicity assessments

In the initial study, total serum antibody titers were measured by enzyme-linked immunosorbent assay (ELISA), and neutralizing serum antibody titers were measured by plaque reduction neutralization test (PRNT). In the initial study, immunogenicity samples were drawn at screening, 2 and 4 weeks after each vaccination, and 6 months after the last vaccination. In the follow-up study, samples were drawn at screening (approximately 2 years after the initial vaccination) for all available participants. Those in the 1×MVA BD and 2×MVA BD groups also had samples drawn at baseline and 1 week, 2 weeks, 4 weeks, and 6 months post-booster.

#### Vaccinia ELISA

Total antibodies were measured using an automated ELISA on a Biomek FX ELISA robot (Beckman Coulter, Germany). 96-well plates were coated overnight with crude antigen preparation (MVA-infected Chicken Embryo Fibroblast lysate) in 200 mM Na_2_CO_3_, pH 9.6. After washing, plates were blocked with PBS, 5% FBS and 0.05% Tween 20. Plates were washed again before the addition of test sera which were titrated in duplicate 2-fold serial dilutions starting at 1:50. After a 1 h incubation, plates were washed, incubated for 1 h with detection antibody (goat anti-human HRP, Sigma-Aldrich) and washed again before a 30-minute development with 3,3’,5,5’-Tetramethylbenzidine substrate. The optical density was measured at 450 nm. The antibody titers were calculated by linear regression and defined as the serum dilution that resulted in the optical density of the assay cut off (0.35).

#### Vaccinia PRNT

Neutralizing antibodies were determined using a PRNT_50_. Test sera were 2-fold serially diluted starting at 1:5 in DMEM, 7% FBS and added to an equal volume of vaccinia virus (Western Reserve strain, ABI, USA), then incubated overnight at 37°C. Following neutralization, the sera were transferred in duplicate to confluent Vero cell monolayers in 48-well cell culture plates. After 70 min adsorption, overlay-medium (DMEM, 7% FBS, Penicillin, Streptomycin, and 0.5% methyl cellulose) was added and then incubated overnight. Following crystal violet staining, plates were scanned and automatically counted. The number of plaques was fitted as a linear function of the log10 of the dilution and the titer expressed as the serum dilution where the virus was neutralized by 50% compared to the 100% virus control (average number of plaques over 40 wells).

### 2.5 Safety Assessments

The safety profile observed for the initial study has been previously reported [29]. Thus, the safety assessments described herein are specific to the follow-up study.

Safety and reactogenicity assessments included solicited local and systemic adverse events, unsolicited adverse events, and serious adverse events. Solicited adverse events constituted a set of predefined, expected local and systemic reactions. Solicited local reactions included erythema, swelling, and induration, which were all graded by size/diameter on a predefined scale of 0 to 3 (0 was absent; 1 was <30 mm; 2 was >30 mm to <100 mm and 3 was ≥100 mm) and pain and pruritus which were also graded on a scale of 0 to 3 (0 was absent; 1 was painful on touch/mild; 2 was painful when limb was moved/moderate; 3 was spontaneously painful/prevents normal activity or severe). Solicited systemic reactions included elevated body temperature, which was graded on a scale of 0 to 4 (0 was <37.5°C; 1 was 37.5°C to <38.0°C; 2 was 38.0°C to <39.0°C; 3 was 39.0°C to 40.0°C; 4 was ≥40.0°C) and headache, myalgia, nausea, and fatigue, which were also graded on a scale of 0 to 4 (0 was none; 1 was mild; 2 was moderate; 3 was severe; 4 was life-threatening or disabling). These solicited adverse events (including the scaling criteria) were listed on a memory aid that was provided to participants for 8 days following the MVA-BN booster vaccination. Unsolicited adverse events were reported at each study visit and consisted of any adverse event that was not listed on the memory aid or occurred outside the 8-day post-booster vaccination period. Local adverse events occurring during the 8-day post-booster vaccination period were all considered related to study vaccination, but the investigator determined causality for all other reported events.

Safety laboratory tests were performed at screening and 2 weeks post-booster vaccination. Abnormal laboratory values that were assessed as clinically significant by the investigator were documented as adverse events. In addition, all Grade 3 (severe) or Grade 4 (potentially life threatening) abnormal values were to be documented as adverse events regardless of clinical significance.

Adverse events of special interest were defined as any clinically significant cardiac symptoms, ECG changes, or cardiac enzymes above normal limits. Participants developing an adverse event of special interest returned for physical or cardiac examinations and further diagnostic tests, as indicated, and were followed-up with until resolution or stabilization.

### 2.6 Statistical methods

Statistical analyses were performed using SAS 9.1 (SAS-Institute, Cary, NC, USA).

The safety and primary immunogenicity analyses were based on the Full Analysis Set (FAS), which comprised all participants who received at least 1 vaccination. Immunogenicity was also summarized using the Per Protocol Set (PPS), which comprised the subset of participants who received all vaccinations and adhered to all protocol conditions without major protocol deviations.

In the initial study, a randomization list was deposited in the eCRF, and in the follow-up study, no randomization or blinding was required. In both studies, conduct was overseen by an independent data and safety monitoring board, and unsolicited adverse events were coded using the most current Medical Dictionary of Regulatory Activities (MedDRA) coding terminology.

The primary immunogenicity variable was the vaccinia-specific seroconversion rate in the initial study and booster response rate (also referred to as seroconversion rate here) in the follow-up study, both based on ELISA. For participants who were seronegative at baseline, seroconversion was defined as the appearance of antibody titers above the limit of detection (≥ 50 for ELISA and ≥ 6 for PRNT) in both studies. For participants who were seropositive at baseline, seroconversion was defined as 2-fold increases compared to the baseline titer in the initial study and as an increase of the antibody titer compared to the booster baseline titer in the follow-up study. For both assays, the geometric mean titer (GMT) was calculated by taking the antilogarithm of the mean of the log_10_ titer transformations. Antibody titers below the detection limit were given an arbitrary value of 1.

## 3. Results

### 3.1 Clinical participant population and conduct of the study

The initial study included a total of 753 participants. Of these, there were 204 participants vaccinated against smallpox in the distant past who were assigned to receive an MVA-BN booster. The other 549 participants, who had never been vaccinated against smallpox, were randomized to receive either placebo, 1 or 2 MVA-BN primary vaccinations (**Figure 1**). Across treatment groups there were 8 participants who did not meet all eligibility criteria and were not dosed or included in the FAS. Primary MVA-BN vaccination was administered to 183 participants in the 2×MVA group and 181 participants in the 1×MVA group; placebo was administered to 181 participants in the PBO group. An MVA-BN booster was administered to 200 participants in the HSPX^+^ group. In total, 745 participants received study vaccination and were included in the FAS.

**Figure 1.**
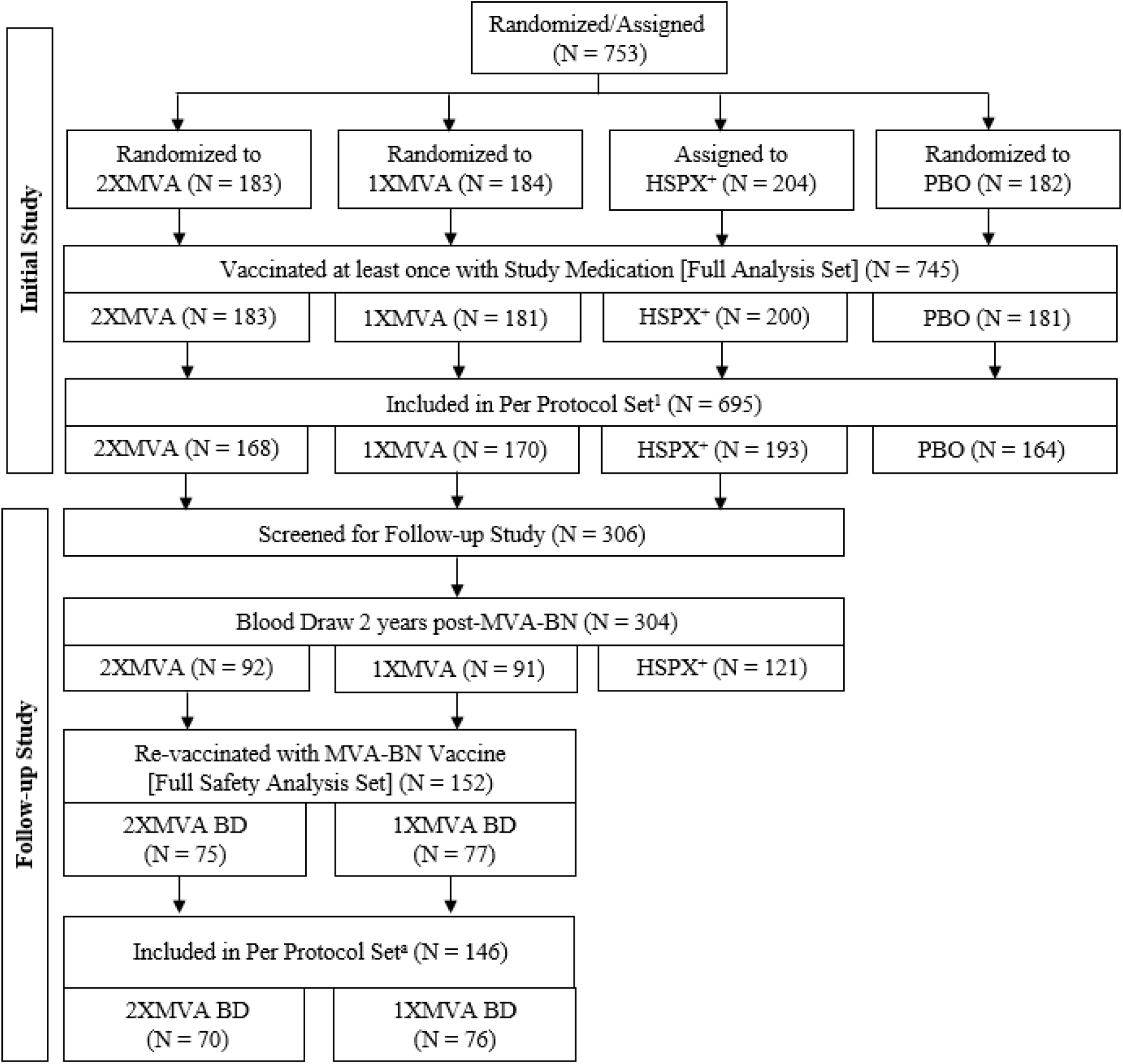
Participant Disposition for Initial and Follow-up MVA-BN Orthopox Vaccination Studies. Abbreviations: BD = booster dose; HSPX^+^ = history of smallpox vaccine positive; N = total number of participants per group; PBO = placebo. ^a^ The Per Protocol Set comprised the subset of participants who received all vaccinations and adhered to all protocol conditions without major protocol deviations.

Approximately 2 years later, 306 participants who received MVA-BN primary vaccinations in the initial study were screened for participation in the follow-up study. Of these, 304 participants (92 from the 2×MVA group, 91 from the 1×MVA group, and 121 from the HSPX+ group) provided blood samples to assess antibody persistence. A total of 75 and 77 participants who initially received 2 or 1 MVA primary vaccinations were revaccinated with an MVA-BN booster and included in the 2×MVA BD and 1×MVA BD groups, respectively.

Study demographic characteristics were generally comparable across treatment groups at the onset of the initial study, with slightly more than half of participants being female (53.0% to 61.9%) and Caucasian (97.2% to 99.0%) (**Table 1**). The HSPX^+^ group had an older mean age (41.5 years) compared with the other treatment groups who were HSPX^−^ (25.3 to 26.0 years), consistent with being old enough to have received a smallpox vaccination prior to worldwide smallpox eradication in 1980. At the initial study baseline, 64.4% of participants were on prior and concomitant medication, with the most common being paracetamol (9.8%), acetylsalicylic acid (7.1%), ibuprofen (5.9%), and ethinyloestradiol (5.0%). All other concomitant medications were reported for less than 2.5% of participants. Similar trends in baseline demographic characteristics (**Table 1**) and prior and concomitant medications were observed in the follow-up study.

**Table 1.** Demographics of Study Participants at Initial and Follow-up Study Baselines.

| **Characteristic** | **2×MVA<sup>a</sup>** | **1×MVA<sup>a</sup>** | **HSPX<sup>+</sup><sup>b</sup>** | **PBO** |
| --- | --- | --- | --- | --- |
| **Initial Study:** | **N = 183** | **N = 181** | **N = 200** | **N = 181** |
| **Age, mean ± SD** |  |  |  |  |
| Years | 25.3 ± 5.0 | 25.4 ± 4.4 | 41.5 ± 7.6 | 26.0 ± 5.1 |
| **Sex, n (%)** |  |  |  |  |
| Female | 97 (53.0) | 112 (61.9) | 115 (57.5) | 107 (59.1) |
| Male | 86 (47.0) | 69 (38.1) | 85 (42.5) | 74 (40.9) |
| **Body Characteristics, mean ± SD** |  |  |  |  |
| BMI, kg/m<sup>2</sup> | 23.6 ± 3.9 | 23.7 ± 3.4 | 24.6 ± 3.9 | 23.9 ± 4.3 |
| **Ethnicity, n (%)** |  |  |  |  |
| Caucasian | 178 (97.3) | 176 (97.2) | 198 (99.0) | 177 (97.8) |
| Asian | 1 (0.5) | 2 (1.1) | 0 | 1 (0.6) |
| Black | 0 | 1 (0.6) | 1 (0.5) | 0 |
| Arabic | 2 (1.1) | 1 (0.6) | 0 | 1 (0.6) |
| Other | 2 (1.1) | 1 (0.6) | 1 (0.5) | 2 (1.1) |
|  | **2×MVA BD<sup>c</sup>** | **1×MVA BD<sup>c</sup>** |  |  |
| **Follow-up Study:** | **N = 75** | **N = 77** |  |  |
| **Age, mean ± SD** |  |  |  |  |
| Years | 27.3 ± 6.0 | 27.9 ± 4.5 |  |  |
| **Sex n (%)** |  |  |  |  |
| Female | 40 (53.3) | 45 (58.4) |  |  |
| Male | 35 (46.7) | 32 (41.6) |  |  |
| **Body Characteristics, mean ± SD** |  |  |  |  |
| BMI, kg/m<sup>2</sup> | 23.7 ± 4.0 | 24.3 ± 3.6 |  |  |
| **Ethnicity, n (%)** |  |  |  |  |
| Caucasian | 74 (98.7) | 74 (96.1) |  |  |
| Asian | 1 (1.3) | 1 (1.3) |  |  |
| Arabic | 0 | 1 (1.3) |  |  |
| Other | 0 | 1 (1.3) |  |  |

Abbreviations: BD = booster dose; BMI = body mass index; n = number of participants; N = total number of participants per group; PBO = placebo; HSPX<sup>+</sup> = history of smallpox vaccination positive; SD = standard deviation; % = percentage based on N.
- <sup>a</sup> Participants in these treatment groups had not been previously immunized against smallpox and received either 1 or 2 primary MVA-BN vaccinations in the initial study. - <sup>b</sup> Participants in this treatment group had been immunized against smallpox in the distant past with replicating smallpox vaccines and received a booster MVA-BN vaccination in the initial study. - <sup>c</sup> Participants in these treatment groups received a booster MVA-BN vaccination 2 years after receiving either 1 or 2 primary MVA-BN vaccinations in the initial study.

### 3.2 Immunogenicity

#### Comparison of Immunogenicity of MVA-BN Booster in Participants Immunized Against Smallpox in the Distant Past with MVA-BN primary vaccination regimens

Following administration of a single primary MVA-BN vaccination in the 1×MVA group, neutralizing antibody (nAb) GMTs rapidly increased relative to initial study baseline at Week 2 (from 1.1 [CI: 1.0, 1.1] to 5.1 [CI: 3.9, 6.6]) and then further increased by Week 4 (7.2 [CI: 5.5, 9.4]) (**Figure 2A and Table 2**). The same trend and magnitude of nAb was recorded in the 2×MVA group following the first MVA-BN vaccination (1.1 [CI: 1.0, 1.2] at baseline, 4.8 [CI: 3.6, 6.3] at Week 2, and then 7.5 [CI: 5.7, 10.0] at Week 4). After receiving the second MVA-BN vaccination at Week 4, nAb GMT in the 2×MVA group further increased and peaked at Week 6 (45.6 [CI: 35.1, 59.3]), nearly 10 times higher than the highest levels observed following the first primary vaccinations. GMT then decreased slightly but remained elevated at Week 8 (34.0 [CI: 26.4, 43.9]).

**Table 2.**
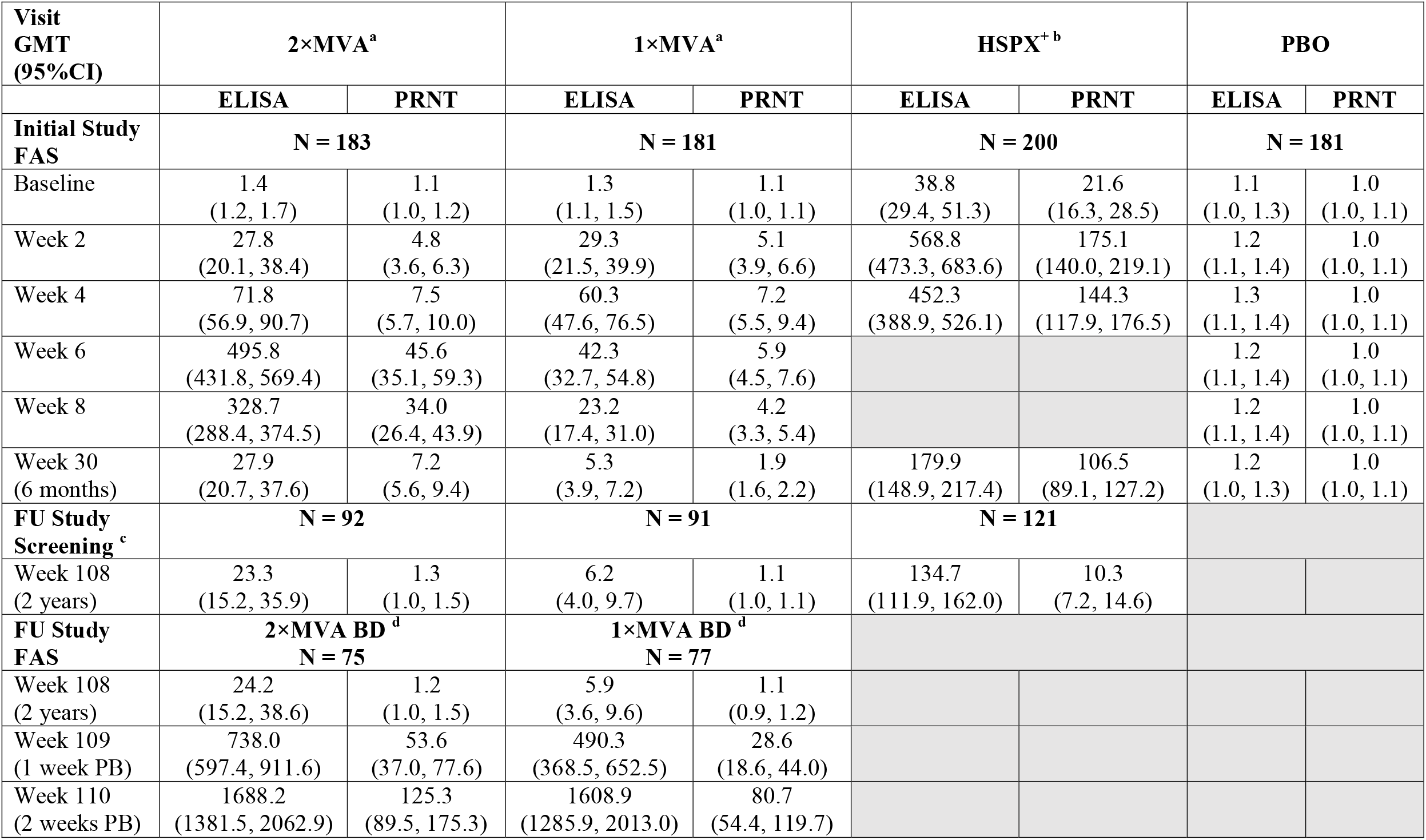
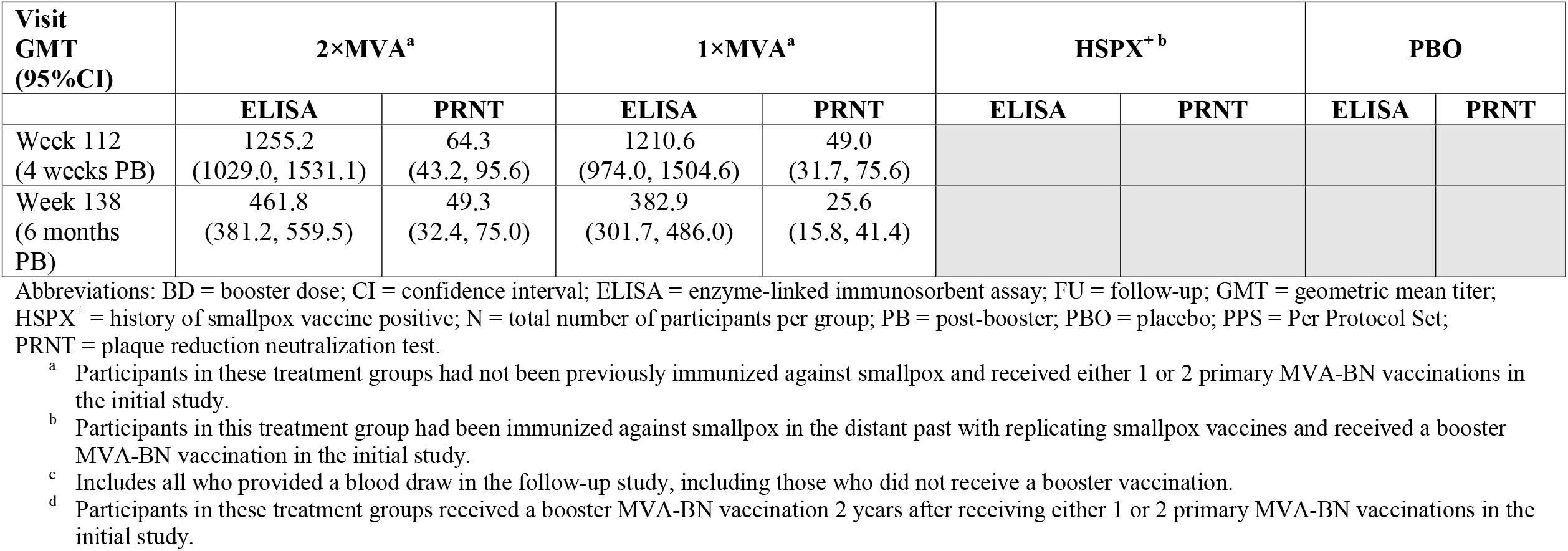
GMTs Over Time in the Initial and Follow-up Studies.

**Figure 2.**
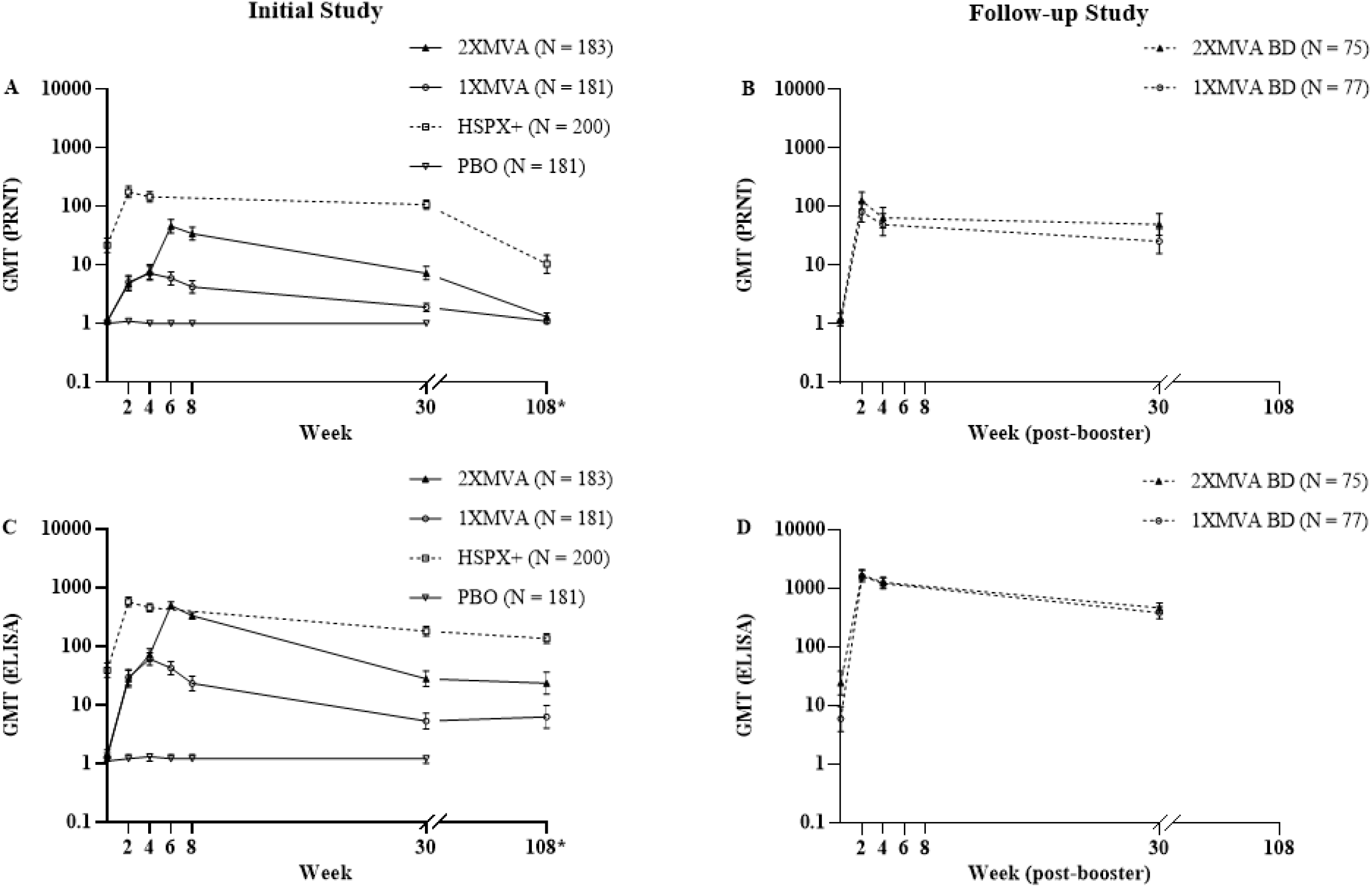
GMTs Over Time in the Initial and Follow-up Studies. Abbreviations: BD = booster dose; ELISA = enzyme-linked immunosorbent assay; GMT = geometric mean titer; HSPX^+^ = history of smallpox vaccine positive; N = total number of participants per group; PBO = placebo; PRNT = plaque reduction neutralization test. Note: Dashed lines are used to connect post-booster datapoints. The 1 week post-vaccination timepoint for the follow-up study (Week 109), included in Table 2, has been omitted from this figure to facilitate comparisons of immune response kinetics between studies. ^*^The 2-year follow-up timepoint (at Week 108) depicts immunogenicity data obtained from all 304 participants in the follow-up study who provided blood draws 2 years post MVA-BN and included 92 participants in the 2×MVA group, 91 participants in the 1×MVA group, and 121 participants in the HSPX^+^ group (for additional details see **Figure 1**).

In the HSPX^+^ group of individuals previously immunized against smallpox, the peak nAb GMT 2 weeks following an MVA-BN booster was approximately 4 times greater than that observed in the 2×MVA group 2 weeks after the second primary MVA-BN vaccination (175.1 [140.0, 219.1] vs 45.6 [35.1, 59.3], respectively). Also in the HSPX^+^ group, nAb GMT remained elevated 4 weeks following the MVA-BN booster (144.3 [CI: 117.9, 176.5]) relative to baseline (21.6 [CI: 16.3, 28.5]) (**Figure 2A and Table 2**).

Approximately half of participants in the 1×MVA and 2×MVA groups seroconverted for nAb at Week 2 (52.0% and 45.1%, respectively), which further increased at Week 4 (62.1% and 56.7%, respectively; **Figure 3A**). The second primary vaccination in the 2×MVA group further elevated seroconversion 2 weeks later at Week 6 (89.2%), remaining stable at Week 8 (86.0%).

**Figure 3.**
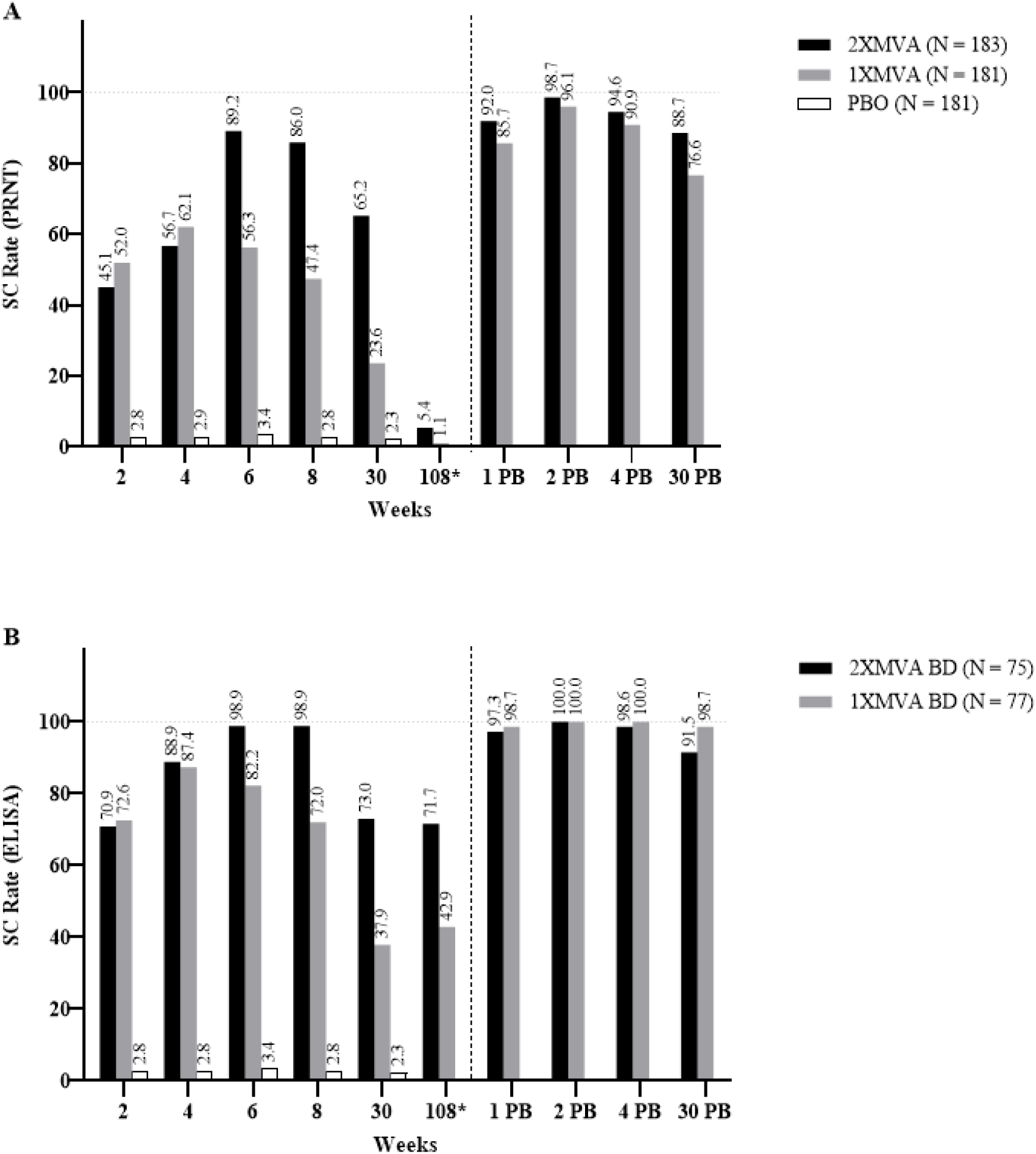
Seroconversion Rates Over Time in the Initial and Follow-up Studies. Abbreviations: BD = booster dose; ELISA = enzyme-linked immunosorbent assay; N = total number of participants per group; PBO = placebo; PRNT = plaque reduction neutralization test; SC = seroconversion. ^*^The 2-year follow-up timepoint (at Week 108) depicts immunogenicity data obtained from participants who provided blood draws 2 years following primary immunization with MVA-BN and included 92 participants in the 2×MVA group and 91 participants in the 1×MVA group (for additional details see **Figure 1**).

The HSPX^+^ group had a wide range of neutralizing antibodies at initial study baseline and therefore very high titers were needed to achieve seroconversion for individuals with higher baseline titers. Nonetheless, 78.5% of the participants were seroconverted at Week 2, before it declined to 69.8% at Week 4, and then 63.8% at 6 months.

The same general post-vaccination trends in neutralizing antibodies, assessed using PRNT, were also observed for total antibodies, assessed using ELISA (**Figure 2C, Figure 3B, and Table 2**). However, while a booster induced higher levels of peak neutralizing antibodies in those vaccinated in the distant past compared to primary vaccination in the 2×MVA group, these levels were more comparable for total antibodies (568.8 [CI: 473.3, 683.6] vs 495.8 [CI: 431.8, 569.4], respectively).

As expected, antibody GMTs and seroconversion rates in the PBO group were low at all timepoints, with negligible changes over the course of the initial study.

#### Immunogenicity at 6 months and 2 Years Following the Last Vaccination

Six months after the last vaccination (at Week 30), nAb GMT remained higher compared with those observed at baseline in the HSPX^+^ group (106.5 [CI: 89.1, 127.2]). Sustained detectable Ab levels above baseline were also observed, although to a lesser extent, in the 1×MVA (1.9 [CI: 1.6, 2.2]) and 2×MVA groups (7.2 [CI: 5.6, 9.4]) (**Figure 2A and Table 2**). Seroconversion based on nAb levels continued to be observed in 23.6% of the 1×MVA group and 65.2% in the 2×MVA group at 6 months (**Figure 3A**).

For participants in the initial study who were assessed for immunogenicity as part of the follow-up study,2-year nAb GMTs were comparable to pre-vaccination levels observed at initial study baseline (**Figure 2A and Table 2**).

When evaluating sustained total antibody levels, GMTs remained elevated at the 2-year timepoint relative to initial study baseline for all groups, with the most notable sustained titers in the HSPX^+^ group (134.7 [CI: 111.9, 162.0] vs 38.8 [CI: 29.4, 51.3]) and more modest sustained titers in the 2×MVA (23.3 [CI: 15.2, 35.9] vs 1.4 [CI: 1.2, 1.7]) and 1×MVA (6.2 [CI: 4.0, 9.7] vs 1.3 [CI: 1.1, 1.5]) groups (**Figure 2C and Table 2)**. The durability of total antibody responses at 2 years was also reflected by sustained ELISA-based seroconversion, similar to that observed at the 6-month timepoint (the last timepoint in the initial study), in the 1×MVA (42.9% vs 37.9%) and 2×MVA (71.7% vs 73.0%) groups (**Figure 3B**).

#### Comparing the Immunogenicity of MVA-BN Booster Vaccinations in Subjects Who Received Primary MVA-BN Vaccination Regimens

Following revaccination with a single MVA-BN booster 2 years after participation in the initial study (Week 108), nAb GMTs rapidly increased 1 week later (Week 109) in both the 1×MVA BD and 2×MVA BD groups (28.6 [CI: 18.6, 44.0] and 53.6 [CI: 37.0, 77.6], respectively). Neutralizing antibodies continued to rise, peaking 2 weeks later (Week 110) in both groups (80.7 [CI: 54.4, 119.7] and 125.3 [CI: 89.5, 175.3], respectively) (**Figure 2B and Table 2**). These booster responses were similar in magnitude and kinetics between treatment groups and exceeded those observed in the initial study. Four weeks post-booster (Week 112) nAb GMTs decreased by nearly half in the 1×MVA BD (49.0 [CI: 31.7, 75.6]) and 2×MVA BD (64.3 [CI: 43.2, 95.6]) groups, although elevated relative to the pre-booster 2-year baseline. At 6 months post-booster, nAb levels remained elevated in the 1×MVA BD (25.6 [CI: 15.8, 41.4]) and 2×MVA BD (49.3 [CI: 32.4, 75.0]) groups and were notably higher than those observed 6 months following the primary vaccinations in the initial study.

Consistent with these results, nearly all participants rapidly underwent PRNT-based seroconversion as early as 1-week post-booster (Week 109) in the 1×MVA BD and 2×MVA BD groups (85.7% and 92.0%). Seroconversion rates peaked 2 weeks post-booster (Week 110: 96.1% and 98.7%) and remained high 6 months later (Week 134: 76.6% and 88.7%) (**Figure 3A**).

Rapid post-booster titer increases that were robust after 1 week and peaked at 2 weeks were also observed for total antibodies assessed by ELISA (**Figure 2D, Figure 3B, and Table 2**). Similar to what was observed with nAbs, peak total antibody GMTs were higher following a booster compared with either 1 or 2 primary vaccinations. Peak post-booster total antibody titers were nearly 3 times higher in the 2×MVA BD group (1688.2 [CI: 1381.5, 2062.9]) compared to the HSPX^+^ group in the initial study (568.8 [CI: 473.3, 683.6]).

Across all timepoints, immunogenicity assessments performed on the PPS were consistent with those summarized for the FAS (data not shown).

### 3.3 Overall Safety Assessment

The safety results of the initial study have been previously described [29]. As such, only the safety results describing participants who were given a 2-year MVA-BN booster vaccination in the follow-up study are described herein.

Approximately half of all follow-up study participants experienced at least 1 unsolicited adverse event (51.3%), and 13.2% of participants experienced vaccine-related adverse events (**Table 3**). Two participants (1.3%), who were both in the 2×MVA group, experienced serious adverse events (gastroenteritis and concussion) that resolved without sequelae and neither was considered related to study medication. A total of 5 participants (3.3%) experienced adverse events of special interest, which included 2 participants in the 2×MVA BD group who experienced individual events of palpitations and 3 participants in the 1×MVA BD group who experienced events of musculoskeletal pain (2 events in 1 participant), palpitations (1 event in 1 participant), and noncardiac chest pain (1 event in 1 participant). All adverse events of special interest were mild or moderate in intensity and considered unrelated to study medication. No deaths occurred during the follow-up study and no participants withdrew or discontinued due to an adverse event.

**Table 3.** Overview of Unsolicited Adverse Events in the Follow-up Study.

| **Parameter**<br>**n (%)** | **2×MVA BD <sup>a</sup>**<br>**N = 75** | **1×MVA BD <sup>a</sup>**<br>**N = 77** | **All Participants**<br>**N = 152** |
| --- | --- | --- | --- |
| **At least one unsolicited:** |  |  |  |
| Adverse event | 40 (53.3) | 38 (49.4) | 78 (51.3) |
| Related <sup>b</sup> AE | 11 (14.7) | 9 (11.7) | 20 (13.2) |
| Serious adverse event | 2 (2.7) | 0 | 2 (1.3) |
| Related serious adverse event <sup>b</sup> | 0 | 0 | 0 |
| Adverse event of special interest | 2 (2.7) | 3 (3.9) | 5 (3.3) |
| Related adverse event of special interest <sup>b</sup> | 0 | 0 | 0 |

Abbreviations: AE = adverse event; BD = booster dose; n = number of participants; N = total number of participants per group; % = percentage based on N.
<sup>a</sup> Participants in these treatment groups received a booster MVA-BN vaccination 2 years after receiving either 1 or 2 primary MVA-BN vaccinations in the initial study.
<sup>b</sup> Definite, probable, possible or unknown relationship to vaccination

Overall, for solicited adverse events, the most commonly experienced local events were injection site erythema (82.2%) and injection site pain (80.3%), while the most commonly experienced systemic events were fatigue (32.2%), myalgia (23.7%), and headache (28.9%) (**Table 4**). The majority of solicited adverse events were mild or moderate in intensity, with the exception of 1 subject in the 1×MVA group and 3 subjects in the 2×MVA group who experienced Grade 3 (severe) events of headache, myalgia, and/or fatigue. No subjects experienced any Grade 4 (life-threatening or disabling) events.

**Table 4.** Overview of Solicited Adverse Events in the Follow-up Study.

| **Event Type**<br>**Preferred Term, n (%)** | **2×MVA BD <sup>a</sup>**<br>**N = 75** | **1×MVA BD <sup>a</sup>**<br>**N = 77** | **All Participants**<br>**N = 152** |
| --- | --- | --- | --- |
| **Local** |  |  |  |
| Injection site erythema | 60 (80.0) | 65 (84.4) | 125 (82.2) |
| Injection site pain | 58 (77.3) | 64 (83.1) | 122 (80.3) |
| Injection site swelling | 51 (68.0) | 48 (62.3) | 99 (65.1) |
| Injection site induration | 58 (77.3) | 58 (75.3) | 116 (76.3) |
| Injection site pruritus | 30 (40.0) | 36 (46.8) | 66 (43.4) |
| **Systemic** |  |  |  |
| Fatigue | 22 (29.3) | 27 (35.1) | 49 (32.2) |
| Myalgia | 17 (22.7) | 19 (24.7) | 36 (23.7) |
| Headache | 19 (25.3) | 25 (32.5) | 44 (28.9) |
| Nausea | 6 (8.0) | 12 (15.6) | 18 (11.8) |
| Body temperature increased | 3 (4.0) | 4 (5.2) | 7 (4.6) |

Abbreviations: BD = booster dose; n = number of participants; N = total number of participants per group;
% = percentage based on N.
<sup>a</sup> Participants in these treatment groups received a booster MVA-BN vaccination 2 years after receiving either 1 or 2 primary MVA-BN vaccinations in the initial study.

Very few related unsolicited adverse events were reported, with injection site warmth being the most commonly experienced (3.9%) (**Table 5**). Pain in extremity, reported by 1 participant in the 2×MVA BD group, was the only severe unsolicited adverse event that was considered possibly related to study treatment. This event was not serious and the participant recovered the same day.

**Table 5.** Overview of Related Unsolicited Adverse Events Experienced by > 1 Participant in the Follow-up Study.

| **Preferred Term**<br>**n (%)** | **2×MVA BD <sup>a</sup>**<br>**N = 75** | **1×MVA BD <sup>a</sup>**<br>**N = 77** | **All Participants**<br>**N = 152** |
| --- | --- | --- | --- |
| Injection site warmth | 3 (4.0) | 3 (3.9) | 6 (3.9) |
| Lymphadenopathy | 3 (3.9) | 0 | 3 (2.0) |
| Chills | 0 | 2 (2.6) | 2 (1.3) |
| Dizziness | 1 (1.3) | 1 (1.3) | 2 (1.3) |
| Injection site irritation | 2 (2.7) | 0 | 2 (1.3) |
| Nasopharyngitis | 2 (2.7) | 0 | 2 (1.3) |

Abbreviations: BD = booster dose; n = number of participants; N = total number of participants per group;
% = percentage based on N.
<sup>a</sup> Participants in these treatment groups received a booster MVA-BN vaccination 2 years after receiving either 1 or 2 primary MVA-BN vaccinations in the initial study.

## Discussion

In this study, an MVA-BN vaccine booster dose induced a robust immune response regardless of smallpox vaccination history and regardless of the use of 1 or 2 doses of MVA-BN at priming:

Two years after either a single or a 2-dose priming in naïve individuals, a rapid increase in neutralizing antibody titers was observed, that was highest at 2 weeks and remained elevated relative to baseline at 6 months after booster vaccination. A single MVA-BN booster administered to those initially vaccinated with older generation replicating smallpox vaccines in the distant past also evoked a rapid increase in neutralizing antibody titers, that was highest at 2 weeks and remained elevated relative to baseline at 6 months after vaccination. The anamnestic neutralizing antibody response was similar in overall magnitude in all groups, although slightly lower in both MVA-BN-primed groups. Booster responses, larger than those seen following primary vaccination, are consistent with what has been previously observed following revaccination with other smallpox vaccines [44].

These data suggest that priming with 1 or 2 doses of MVA-BN can induce B-cell immune memory similarly to that of older generation replicating smallpox vaccines, which have conferred long-term protection and are believed have prevented orthopox outbreaks (including Monkeypox transmission) until recently [7-9].

While neutralizing antibodies in those immunized long ago were detectable decades later, a protective nAb threshold has not been defined for smallpox, monkeypox, or other orthopox viruses. At booster study baseline, those who received primary MVA-BN immunizations 2 years earlier exhibited detectable nAb levels that had declined to near baseline levels. Yet, even with low levels of neutralizing antibodies 2 years later, a single MVA-BN booster induced robust anamnestic responses regardless of whether a 1- or 2-dose primary vaccination schedule had been administered. These responses peaked after only 2 weeks and exhibited the same rapid kinetics as those who received an MVA-BN booster after being immunized long ago with an older generation replicating smallpox vaccine. This demonstration of long-lived B-cell memory strongly supports protection in the absence of persisting neutralizing antibodies, particularly for diseases like smallpox, with longer incubation periods [45].

Sustained MVA-BN-induced protection has been observed in nonhuman primates challenged with monkeypox virus [46, 47]. Although post-immunization neutralizing antibody titers fell to pre-vaccination levels after about three years, antibody levels quickly rose upon exposure to the virus and the animals were protected against manifestation of the disease. This provides evidence that MVA-BN vaccination is not only capable of inducing a robust anamnestic response, but that this response can be associated with disease protection.

This type of long-term immunological memory, observed in the absence of sustained neutralizing antibodies, has been demonstrated with vaccines for hepatitis B virus (HBV) and measles, mumps, and rubella (MMR). In HBV studies, 90% to 100% of previously vaccinated participants who were seronegative exhibited rapid anamnestic responses when given a booster vaccination [48, 49]. Neutralizing mumps antibodies were found to decrease over time [50], with nearly half of young adults who had received MMR vaccinations 20 years earlier as infants being seronegative. However, over 70% of seronegative individuals had an anamnestic response upon revaccination [51].

In this study, MVA-BN induced robust total antibody responses in all groups, which was proportionally higher as compared to the neutralizing antibodies in those who had been primed with 1 or 2 doses of MVA-BN than those who had been primed with older generation replicating vaccines in the distant past. Although the difference in time between priming and booster in the different groups may be a confounding factor, increasing evidence suggests that non-neutralizing antibodies may play significant roles in protecting against some viral infections [52, 53]. These non-neutralizing antibodies may elicit effector pathways such as antibody-dependent cellular cytotoxicity (ADCC), antibody-dependent cellular phagocytosis (ADCP), and antibody-mediated complement-dependent cytotoxicity (CDC). Further, less specific antibodies can target less variable, cross-serotype viral proteins in addition to the targeted protein, while the neutralizing antibodies are primarily effective against the targeted protein. This extremely high total antibody response, induced by MVA-BN, may be important in providing cross-protection against other orthopox viruses such as monkeypox [46, 47].

No safety concerns were identified in the follow-up study, for the subset of participants administered a booster dose of MVA-BN 2 years following primary vaccination. This is consistent with the safety profile previously reported for the initial study [29]. Since reports of myopericarditis were reported in military personnel administered replicating smallpox vaccines [13, 16, 54, 55], cardiac events have been intensively monitored throughout the MVA-BN clinical development program. Although 5 participants experienced cardiac adverse events of special interest that included palpitations, musculoskeletal chest pain, and non-cardiac chest pain, all of these events were mild or moderate in intensity and considered unrelated to study medication. Thus, the safety findings from this study further support the known safety profile of MVA-BN in adults, which has not identified any cardiac safety concerns or confirmed cases of myo- or pericarditis [27, 29, 30].

## 5 Conclusion

Priming with either 1 or 2 doses of MVA-BN induced a durable immune memory, similar to that of older generation replicating smallpox vaccines. When the MVA-BN vaccine was administered as a booster to individuals either primed 2 years earlier with MVA-BN or an older generation replicating smallpox vaccine in the distant past, it elicited a similar rapid and durable immune response that was generally safe and well-tolerated. A one-dose priming with MVA induced a long-term B-cell memory that resulted in robust anamnestic responses following an MVA booster 2 years later, suggesting that a single dose may offer protection against future exposure.

## Data Availability

All clinical trial results are available online at:
https://clinicaltrials.gov/ct2/show/results/NCT00316524?term=NCT00316524&draw=2&rank=1
https://clinicaltrials.gov/ct2/show/results/NCT00686582?term=NCT00316524&draw=2&rank=2

## Acknowledgements

The study described herein was sponsored by Bavarian Nordic and funded by the National Institute of Allergy and Infectious Diseases (N01-AI40072). The investigators wish to thank all study participants. We would like to acknowledge in memoriam the contributions of Frank von Sonnenburg and Stephan de la Motte. We also thank Josef Weigl, Alfred v. Kremplhuber, Siegfried Rösch, and Garth Virgin for their efforts.

## Author Contribution Statement

**Heiko Ilchmann:** Conceptualization, Supervision, Investigation (trial site), Writing – review and editing; **Nathaly Samy:** Conceptualization, Supervision, Writing – review and editing; **Daniela Reichhardt**: Project administration, Writing – review and editing; **Darja Schmidt**: Supervision, Investigation (laboratory), Validation, Writing – review and editing; **Jacqueline D Powell**: Writing – Initial draft preparation, Writing – review and editing; **Thomas PH Meyer**: Supervision, Investigation (laboratory), Validation, Writing - review and editing, Visualization, Validation; **Günter Silbernagl**: Formal analysis, Validation, Software, Writing – review and editing; **Rick Nichols:** Supervision, Investigation (laboratory), Validation, Writing – review and editing; **Laurence De Moerlooze:** Supervision, Writing – review and editing; **Liddy Chen:** Formal analysis, Writing – review and editing; **Heinz Weidenthaler**: Conceptualization, Methodology, Supervision, Writing – review and editing; **Paul Chaplin**: Conceptualization, Methodology, Supervision, Funding acquisition, Writing – review and editing.

All authors attest they meet the ICMJE criteria for authorship.

